# Genome-wide study of half a million individuals with major depression identifies 697 independent associations, infers causal neuronal subtypes and biological targets for novel pharmacotherapies

**DOI:** 10.1101/2024.04.29.24306535

**Authors:** Andrew M McIntosh, Cathryn M Lewis, Mark J Adams for the Psychiatric Genomics Consortium Major Depressive Disorder Working Group

## Abstract

In a genome-wide association study (GWAS) of 685,808 individuals with major depression (MD) and 4,364,225 controls from 29 countries and across diverse and admixed ancestries, we identify 697 independent associations at 636 genetic loci, 293 of which are novel. Using fine-mapping and functional genomic datasets, we find 308 high-confidence gene associations and enrichment of postsynaptic density and receptor clustering. Leveraging new single-cell gene expression data, we conducted a causal neural cell type enrichment analysis that implicated excitatory and inhibitory midbrain and forebrain neurons, peptidergic neurons, and medium spiny neurons in MD. Critically, our findings are enriched for the targets of antidepressants and provide potential antidepressant repurposing opportunities (e.g., pregabalin and modafinil). Polygenic scores (PGS) from European ancestries explained up to 5.7% of the variance in liability to MD in European samples and PGS trained using either European or multi-ancestry data significantly predicted case control status across all four diverse ancestries. These findings represent a major advance in our understanding of MD across global populations. We provide evidence that MD GWAS reveals known and novel biological targets that may be used to target and develop pharmacotherapies addressing the considerable unmet need for effective treatment.

## Introduction

Major depression (MD) is a leading cause of worldwide disability and affects approximately 15% of the population during their lifetime. The peak age of onset is in early adulthood and the disorder is typically recurrent or chronic in nature, with persisting disability despite pharmacological and psychological therapies. Twin and family-based studies provide evidence of a significant genetic contribution to its etiology, with a heritability of approximately 37% (Polderman et al., 2015). Since 2013, genome-wide association studies (GWAS) have provided major insights into the polygenic nature of MD, its genetic risk factors and underlying mechanisms (Ripke et al., 2013; CONVERGE Consortium et al., 2015; Hyde et al., 2016; Howard et al., 2018; Wray et al., 2018; Howard et al., 2019; Levey et al., 2021; Als et al., 2022; Meng et al., 2024). The largest study conducted to date involved a meta-analysis of the Million Veteran Program, 23andMe, UK Biobank, FinnGen, and iPSYCH including 371K cases (Als *et al*., 2022) reporting 243 independent MD risk loci.

Nevertheless, the molecular, cellular, and neurobiological mechanisms of MD remain largely unidentified, limiting the development of disease models and mechanism-informed drug treatments (Zhu, 2020). In the current study, we report results from the Psychiatric Genomics Consortium (PGC) Major Depressive Disorder Working Group’s largest GWAS of MD to date. We used strategies designed for analysis of multi-ancestry and admixed populations to implement the largest, most inclusive study of MD genetics to date. These results substantially extend previous GWAS findings, provide evidence of causal genes, cell types and tissues involved in MD, and demonstrate out-of-sample prediction across diverse ancestry groups.

## Results

### GWAS associations

We meta-analyzed (see **Methods** and **Figure S1, Tables S1, S2, supplementary study information**) GWAS summary statistics from 109 ancestrally diverse cohort datasets with 685,808 cases and 4,364,225 controls (with power equivalent to a case-control study of 1,000,101 cases and 1,000,101 controls, with 23% in diverse/non-European ancestries, Table 1). For cohorts with diverse ancestries, associations were assessed using tools that explicitly model population structure, admixture, and relatedness (GENESIS). For a subset of cohorts with ancestrally diverse samples, we compared the sample size using the commonly used strategy of assigning individuals into ancestry groups followed by logistic regression (N=24,859) with our joint approach (N=47,642) and found a 92% sample size increase, leading to a final sample size of 160,611 cases and 1,001,890 controls (Supplementary Material) and the discovery of an additional 66 genome wide significant loci. Using conditional and joint GCTA-COJO (Yang et al., 2012) analysis with threshold P ≤ 5 × 10^-8^ within 10 Mb windows, we found 697 significant independent SNPs in 635 genomic regions, about half (293/635; 46%) of which were novel MD associations (Figure 1, Table S3). Of these, 27 were identified due to the inclusion of cohorts with ancestrally diverse samples.

**Figure 1:**
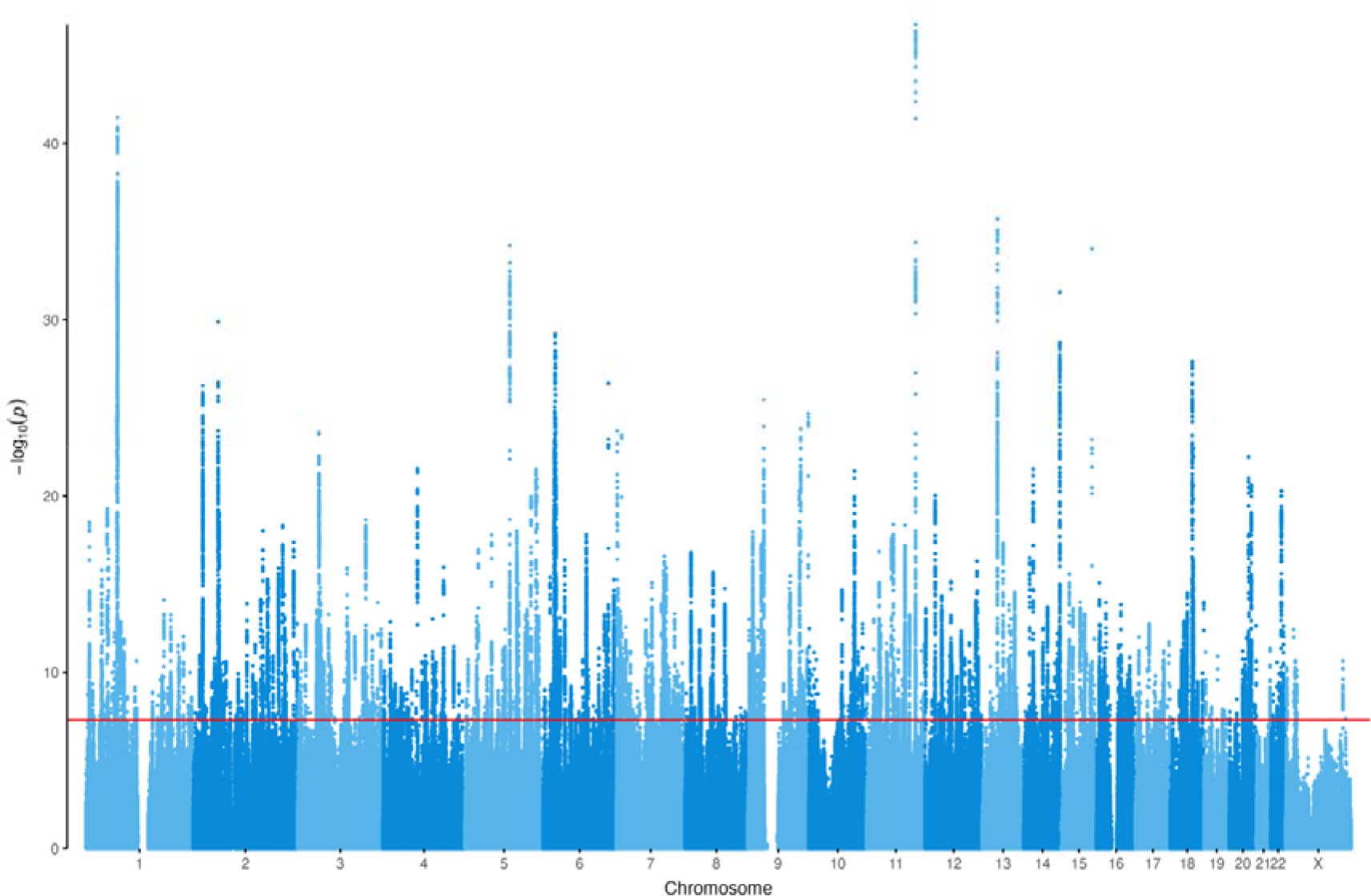
Manhattan Plot of GWAS meta-analysis of 685,808 MD cases and 4,364,225 controls ***Legend****: Manhattan plot displaying the significance of each SNP’s association with MD across the genome (vertical axis). Chromosomal position is shown on the horizontal axis. The horizontal line at 7.3 (-log_10_(5 x 10^-8^)) depicts the statistical significance threshold*.

**Table 1:** Details of diverse ancestry studies included in the current GWAS Legend to. table 1: Table shows the breakdown of the studies included in the current meta- analysis broken down by ancestry group and admixture/multiple ancestries (analysed using GENESIS software). The complete dataset included 109 individual datasets from 93 individual studies, some of which (e.g. UK Biobank) contributed more than one non-overlapping ancestry specific dataset. Neff/2 represents the equal case and control sample sizes of an equivalently powered balanced study.

| Ancestry Group | N studies | N cases | N controls | Neff/2 | N GWAS hits |
| --- | --- | --- | --- | --- | --- |
| European | 76 | 525,197 | 3,362,335 | 788,603 | 570 |
| East Asian | 7 | 18,709 | 349,619 | 30,654 | 0 |
| South Asian | 1 | 3,748 | 25,934 | 6,549 | 0 |
| African | 8 | 9,649 | 122,347 | 17,077 | 0 |
| Hispanic/Latin American | 5 | 19,927 | 340,403 | 36,875 | 0 |
| Multiple | 12 | 108,578 | 163,587 | 120,342 | 0 |
| All ancestries | 109 | 685,808 | 4,364,225 | 1,000,101 | 636 |

**Table 2:**
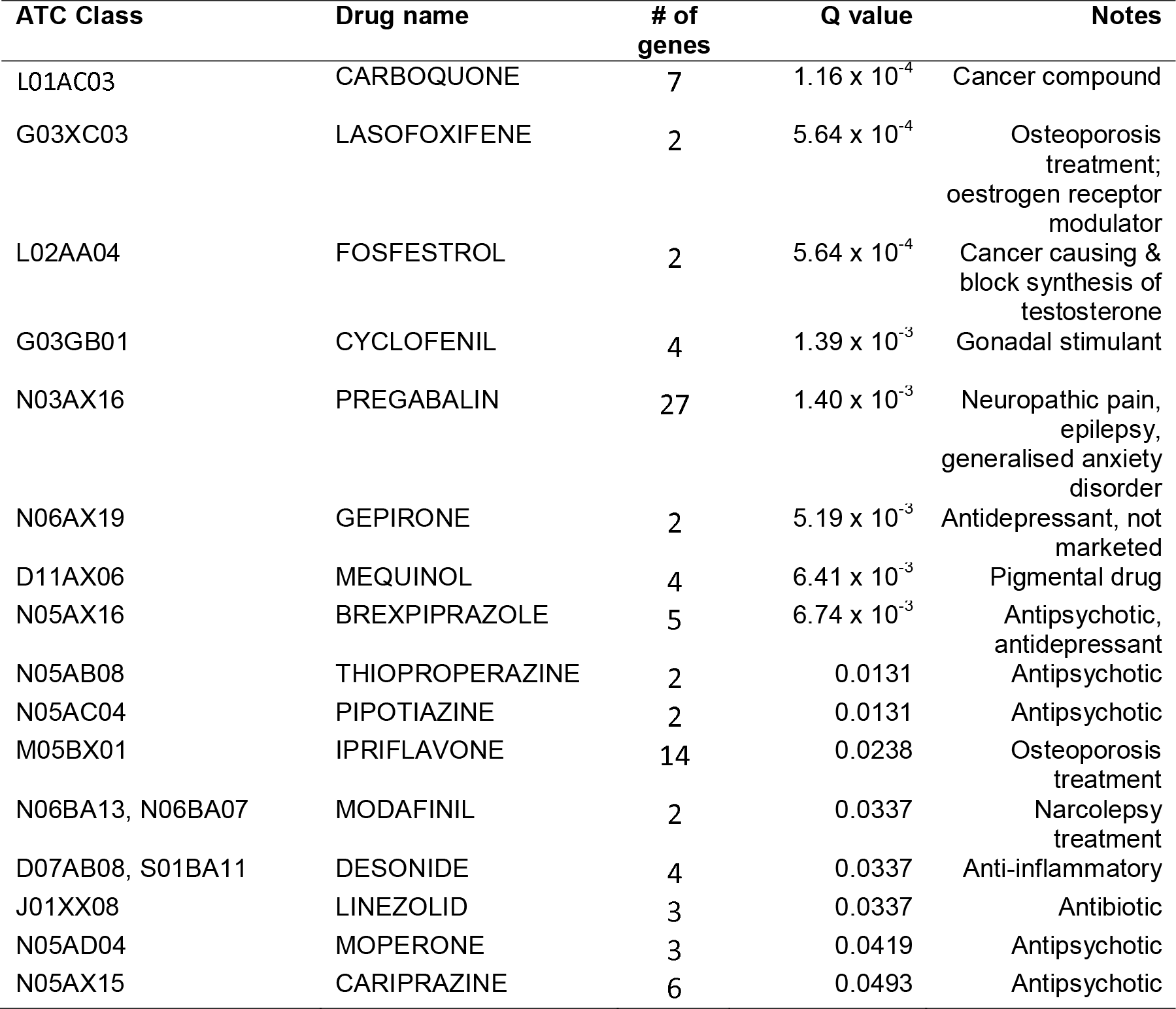
Significant drug target enrichments *Legend*. *Table shows the top 16 most significantly enriched drugs based on capture of their targets within the gene-based associations of the current MD GWAS analysis. One topical preparation is not shown. The test for drug enrichment is not directional and may indicate compounds that confer risk of MD or exacerbate depressive symptoms, as well as those that ameliorate risk or depressive symptoms. Q value is false discovery rate, Benjamini–Yekutieli corrected (competitive analysis)*.

We carried out a fixed-effects meta-analysis for samples of European ancestries (525,197 cases and 3,362,335 controls), for which we had a large single linkage disequilibrium reference dataset, to carry out downstream analyses including heritability, gene prioritization, enrichment, genetic correlation and Mendelian Randomization analyses To examine the consequences of MD phenotyping on the meta-analyses, we implemented genomic structural equation modelling (SEM) with a common factor meta-analysis of the European-ancestry summary statistics in Genomic SEM (Grotzinger et al., 2019) (**Figure S2**). Cohorts were first meta-analyzed based on how the MD phenotype was determined: clinical/interview, electronic health record [EHR], questionnaire, or self-report of MD diagnosis. The proportion of total effective sample size contributed by each phenotype definition was 4% clinical/interview, 54% EHR, 14% questionnaire, and 27% self-report. The different phenotype definitions of MD had strong genetic correlations (LD score *r_g_* from 0.78 to 0.88). We fit a common factor model in Genomic SEM and treated the clinical/interview phenotype as the primary phenotype by fixing its factor loading to 1 and its residual variance to 0. This factor model was consistent with the data (X^2^ = 4.49, P = 0.213), meaning that we could not reject null hypothesis that a single factor capturing all the variance of the primary phenotype explained the intercorrelations among the other depression phenotypes. Most MD phenotypes had strong positive loadings on the common factor (clinical/interview = 1.0 [reference], EHR = 0.92±0.04, questionnaire = 0.95±0.04) although the loading for self-reported diagnosis was lower (self-report loading = 0.85±04). One locus showed significant SNP heterogeneity between phenotyping definitions (rs12124523 intronic variant in *NEGR1*, common factor association P = 8.4 × 10^-14^, Q heterogeneity P = 2.9 × 10^-10^, *I^2^* = 0.71) with a stronger association found in self-reported depression studies (Self-report odds ratio (OR) = 1.081, CI = 1.065–1.098, other cohorts OR = 1.008, CI = 0.999–1.018). We found no evidence of heterogeneity at 569/570 loci, supporting the use of multiple phenotypes in genetic association studies of MD.

SNP-based heritability was estimated in European ancestries using SBayesS (Zeng et al., 2021) to be 8.4% (s.e. 0.07%) on the liability scale (assuming lifetime MD risk of 15%) similar to prior estimates (Howard *et al*., 2018; Wray *et al*., 2018). SBayesS provides genetic architecture estimates of polygenicity of 6% and selection parameter of -0.54. Compared to previously reported estimates for 155 traits, MD has a relatively higher polygenicity, but its associated variants are under weaker negative selection (Zeng *et al*., 2021).

### Gene prioritization and pathway enrichment analysis

We used a range of methods and functional genomic datasets to gain insight into the variants, genes, and pathways that may underlie MD-associated loci: SNP-based fine-mapping of MD-associated loci, integration of expression and protein quantitative trait loci (eQTL and pQTL) data, transcriptome- and proteome-wide association studies (TWAS and PWAS) with summary data-based Mendelian randomization (SMR), colocalization (COLOC) and TWAS-based fine-mapping (of eQTLs and pQTLs, in FOCUS) analyses. We also mapped associated loci to genes using gene-based association analysis in fastBAT, chromatin interaction datasets (HiC) and applied a novel gene prioritization package PsyOPS (**see Methods**).

We undertook functionally informed SNP-based fine-mapping analyses, using the European ancestry GWAS findings, targeted at all autosomal GWAS loci excluding the Major Histocompatibility Complex (MHC) region. Twenty-four variants showed strong putative evidence of causality (posterior inclusion probability, PIP > 0.95, **Table S4**). Credible causal set sizes comprising ≤10 variants (cumulative PIP > 0.95) were identified at 224 loci (**Figure S3**) and 234/564 autosomal loci could be mapped to one or more genes (**Table S5**).

Both eQTL and pQTL data were used to infer up- or down-regulated gene expression or protein levels associated with MD. Stringent criteria were used to identify high confidence associations with MD (**Methods**). MD genetic associations were found to correlate and colocalize with cis-regulated expression of 75 genes (**Table S6**) and cis-regulated levels of 10 proteins (**Table S7**). No gene was identified as high confidence by both eQTL and pQTL analyses.

In total, across SNP-based fine-mapping, eQTL and pQTL analyses, 308 high-confidence associations were identified, with 14 eQTL genes and 1 pQTL gene also identified as high confidence by SNP-based fine-mapping. For example, SNP-based fine-mapping found all SNPs in one 95% credible set were within the cytochrome P450 gene *CYP7B1,* which was also inferred to have decreased expression in the dorsolateral prefrontal cortex of individuals with MD (TWAS p-value = 2.92x10^-15^, COLOC PP4 = 0.939, FOCUS PIP = 1).

Positional mapping approaches were used to identify additional genes that may be involved in MD etiology, including identification of the nearest gene to lead MD variants, aggregating genetic associations across gene regions using fastBAT (**Table S8, Figure S4**), and linking associated loci to genes through Hi-C chromatin interactions using H-MAGMA (**Table S9**). Furthermore, the gene prioritization method PsyOPS was used to score genes based on prior information on mutational constraint, brain expression, and involvement in neurodevelopmental disorders (**Table S10**). Of the 18,737 genes assessed using fastBAT, 1,568 were associated with MD (*P* < 2.67 x 10^-6^) with the dopamine receptor D2 (*DRD2*) gene providing the strongest association (P = 9.39 × 10^-47^). *DRD2* was also identified as associated with MD by H-MAGMA with astrocyte cells providing the strongest evidence from the four brain tissue profiles analyzed (P = 2.72 × 10^-15^). An additional 1,033 genes were also identified as associated with MD based on three-dimensional chromatin data using H-MAGMA. While PsyOPS prioritized a neighboring gene, *NCAM1* (PsyOPS score = 0.402), *DRD2* had an equivalent score (0.399). Other genes with high PsyOPS prioritization scores were *PTPRT, SLC12A5, RFX3, ELAVL2, HCN1, KIF5A, and SHANK3*.

### Synaptic gene set enrichment

We used the high-confidence gene list from SNP-based fine-mapping, TWAS, and PWAS to identify enriched synapse functions using SynGO (Koopmans et al., 2019). The 43 genes from the high-confidence gene list with SynGO annotations were compared against a background of 18,035 brain-expressed genes. We replicated earlier findings from Howard et al (2019), showing enrichment of neuron differentiation processes and postsynaptic membrane components. The current GWAS provided greatly increased specificity, implicating the cytosol, active zone membrane, calcium levels, vesicle cycle and presynaptic endocytosis. Postsynaptically, there was enrichment of synaptic specialization, density, and receptor clustering (**Table S11A+B**).

### Tissue and cell type enrichment analysis

We conducted tissue and cell type enrichment analysis using published expression datasets including bulk RNA-sequencing data from human tissues (Bryois et al., 2020) and single-cell RNA-sequencing data from the adult mouse central and peripheral nervous system (Zeisel et al., 2018). Across human tissues, we found clearer enrichment patterns of MD SNP-heritability in neural tissues using the current GWAS association findings than those obtained from the previous two PGC MDD group analyses **(Figure S5)**. In the adult mouse central and peripheral nervous system, we found significant enrichment of MD SNP-heritability in 10 out of 39 cell types with two different methods (MAGMA and partitioned LD Score, see **Figure 2 and Figure S6**). We confirmed all the cell types identified in the previous GWAS (Wray *et al*., 2018) including both excitatory and inhibitory neurons, but implicate multiple additional inhibitory neuron categories and peptidergic neurons.

**Figure 2:**
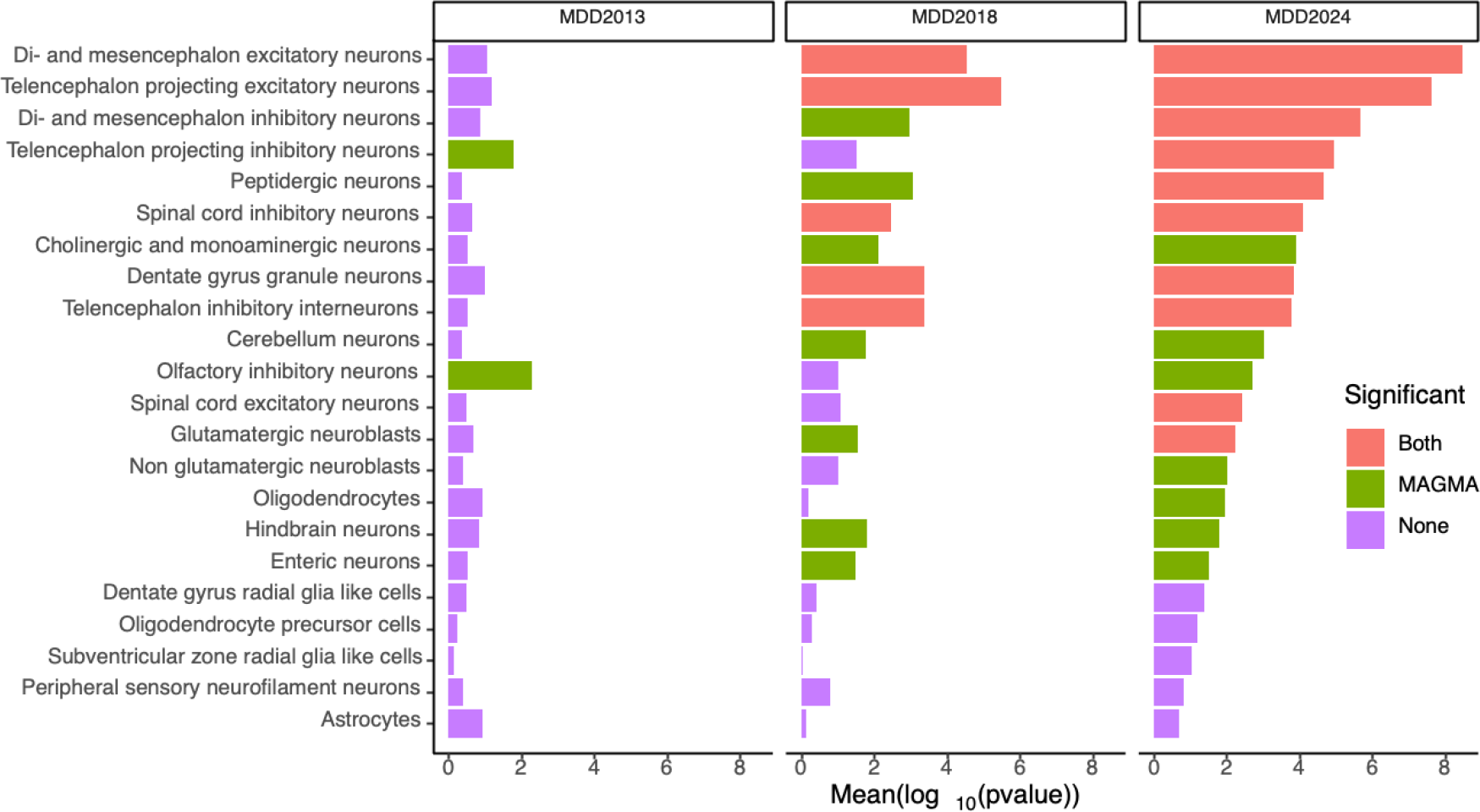
Broad brain cell category enrichment analysis**. Legend to** Figure 2**: Cell type enrichment analysis**. *20 categories of brain cell types are listed (from a total of 39 broad cell type categories tested) along the vertical axis, horizontal bar size represents the significance of the enrichment measured using MAGMA gene set enrichment test or partitioned LDSC. Bars drawn in salmon color represent enrichments that were significant using both methods, green – MAGMA only, blue partitioned LDSC only and purple when neither method showed significant enrichment. 19 broad categories not displayed were also not significant using either method. Columns represent the results of each test using the summary statistics from MDD2013 (Ripke et al., 2013), MDD2018 (Wray et al., 2018) and the current study*.

Analysis at a more refined level of cell types further emphasized the enrichment in excitatory and inhibitory neurons in multiple brain regions (**Figure S6**, **Table S12**). Associated cell types using both methods included midbrain (mouse atlas reference: MEGLU7, MEGLU8, MEGLU10 and MEGLU11), amygdala (TEGLU22), hippocampal (CA1, TEGLU21), thalamic (DEGLU4) and cortical (TEGLU1, TEGLU4, TEGLU8, TEGLU8, TEGLU11, TEGLU13 and TEGLU20) excitatory neurons. We also found additional evidence for the involvement of D1/D2 midbrain and striatal medium spiny neurons (MSN2 by both methods and MSN1,3-4 by MAGMA only).

### Drug target enrichment analysis

Using Drug Targetor, we searched for Anatomical Therapeutic Chemical (ATC) drug classes whose targets were enriched in the signals from the GWAS analysis (Gaspar et al., 2019). Drug Targetor harnesses drug bioactivity data, to prioritize drugs and targets for a given phenotype. Replicating an earlier analysis, we found the gene targets of antidepressants (ATC class N06A) are significantly enriched (**Figure S7**) in our association findings. Other drug classes that were significantly enriched included antipsychotics (N05A), which includes some medicines with antidepressant effects (**Table S13B**).

The gene targets of *specific drugs* were also enriched in genetic associations with MD, although the analysis does not infer whether the effects of these agents were more likely to be congruent or opposed to the effects of genetic risk. The identified drugs provide possible repurposing opportunities and examples included several anti-cancer therapies, and the agents pregabalin (used in the management of pain and anxiety) and modafinil which is used to treat daytime sleepiness caused by narcolepsy (**Tables 2 and S13A**).

### Within and cross-trait prediction

#### Polygenic score prediction in European ancestry samples

Using the case-control cohorts in the meta-analysis, we conducted a leave-one-cohort-out GWAS meta-analysis for 43 European ancestry cohorts that had provided individual level data. Polygenic scores (PGS) were generated in the left-out target European samples generated using SNP weights for the multi-ancestry and the European ancestry meta-analyses derived using SBayesR (Lloyd-Jones et al., 2019). Other PGS methods including the standard p-value clumping and thresholding gave similar results (**Table S14**). Across all European cohorts, the variance explained on the liability scale (was 5.8% (s.e. 0.2%) (**Figure S8, S9)**, with an AUC statistic of 0.625 (**Figure S10**). Adding functional annotations into the algorithm to generate SNP weights for PGS (SBayesR) increased prediction accuracy by 0.1% (i.e., of 5.9%). The was more than 1.4 times greater than that reported in the PGC MDD 2018 analysis (Wray *et al*., 2018; Ni et al., 2021) (**Figure 3a and S8**). The OR for being a case per standard deviation (SD) increase in PGS was 1.57. The OR for being a case in the tenth compared to the first decile of polygenic scores was 4.92 (95% CI 4.57-5.29) (**Figure 3**), and the OR for the top vs bottom centiles was 11.8 (95% CI 8.4-15.2) (**Figure S11**). The non-linear shape of these decile and centile plots is expected under a polygenic architecture (Baselmans et al., 2021). Heterogeneity in the out-of-sample prediction results could be partly explained by the recorded ascertainment type **(Figure 3b and S9),** which we classified as ‘clinical’ (16 cohorts; ascertained from in- or out-patient settings, or EHR) or ‘community’ ascertained (32 cohorts; interviews or questionnaires self-reporting on lifetime depression). The difference in mean PGS between clinical vs community cases was 0.131 (s.e 0.012, P< 2x10^-16^) control sample SD units.

**Figure 3:**
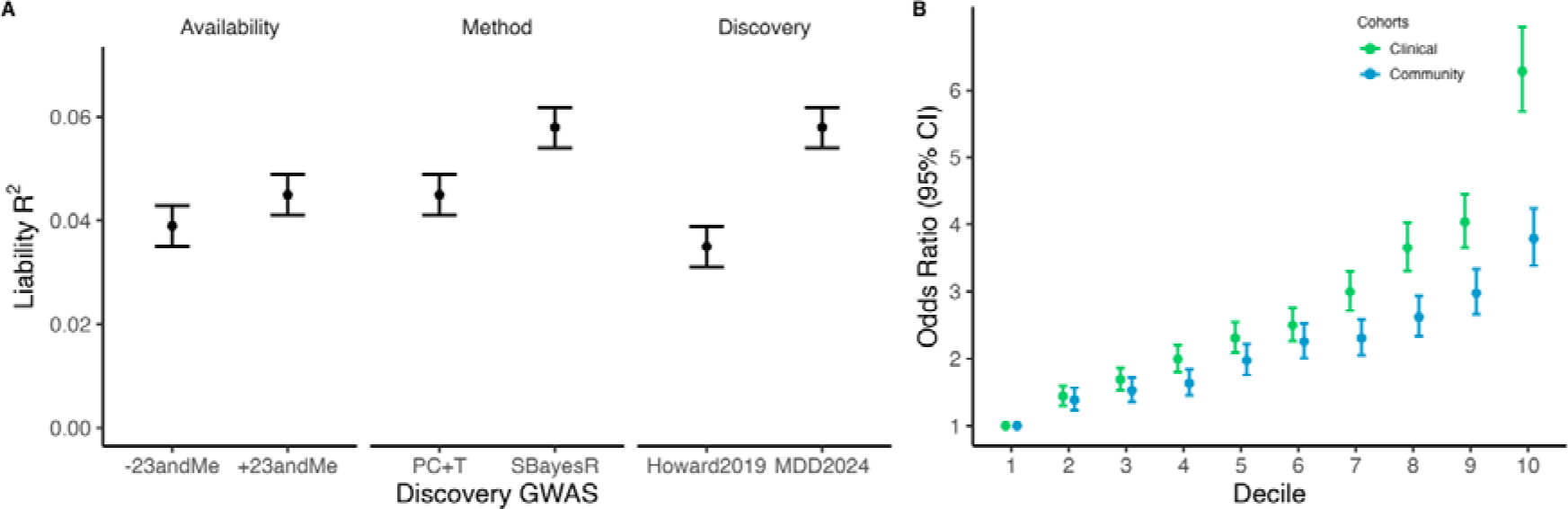
MD polygenic score prediction into European ancestry studies ***Legend: (A)*** *Comparison of liability R^2^ by input summary statistics by availability (full dataset with 23andMe versus public dataset without 23andMe, using P-value clumping + thresholding at P <= 0.05 [P+CT]), PGS method (P+CT versus SBayesR), and discovery dataset (previous Howard 2019 versus current MDD2024 SBayesR). The R2 are estimated across 43 cohorts with individual level data when each was left out of the discovery sample in turn. For the Discovery panel the R2 are estimated from the 20 cohorts with individual level data contributed to the PGC subsequent to the Howard 2019 study. The* r^2^ *was calculated using a lifetime prevalence of 0.15. **(C)** Odds ratio by decile, with reference to decile 1, for clinical and community ascertained studies (SBayesR). Bars reflecting the 95%CI are based on estimates from the logistic regression*.

### Cross-ancestry prediction of MD

We used data from 9 diverse ancestry studies to assess PGS transferability (**Table S15**) using PGS derived from the clumping and thresholding approach. The PGS were derived from the multi-ancestry and the European meta-analysis excluding 23andMe because of data access restrictions (N_effective_ = 739,180 and 576,327, respectively) **(Figure 4, Table S16**). In the diverse ancestry studies, the r^2^, by the PGS based on the European ancestry training data ranged from ∼0.6-4.5%. The r^2^ values for prediction into European ancestry (excluding 23andMe) were 3.9% (s.e. 0.2%) using P_T_=0.05. Values were lowest in studies with participants of African descent, and in the largest African ancestry study, the Million Veteran Program (MVP), the PGS was not associated with MD r^2^=0.0018). Results using the multi-ancestry summary statistics showed only minor and non-significant differences from European-only PGS GWAS trained scores in all ancestry groups.

**Figure 4:**
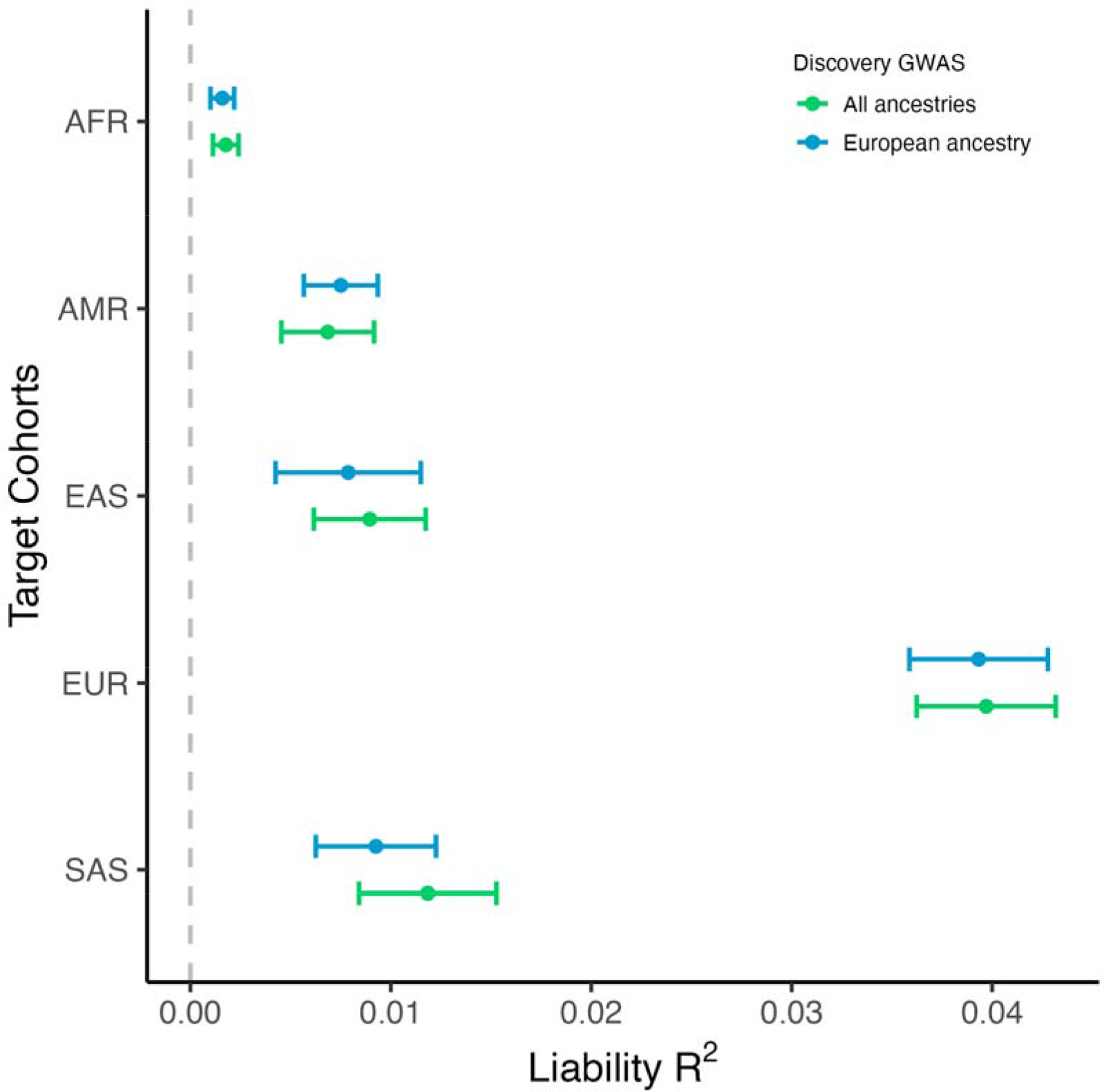
Polygenic prediction of MD status from European and multi-ancestry GWAS into ancestrally diverse non-European studies ***Legend:*** *Details of cohorts found in Table S2. The was calculated using a prevalence of 0.15 with the P+CT method. The error bars are confidence intervals calculated using bootstrap. The training data did not include 23andMe, because of access limitations. AFR: African ancestry; AMR: Hispanic and Latin American ethnicities; EAS: Easy Asian ancestries; EUR: European ancestries; SAS: South Asian ancestries*.

### Genetic correlation and phenome-wide Mendelian randomization analysis

We estimated LD score regression genetic correlations using 3,229 sets of summary statistics obtained from the OpenGWAS catalog (**Table S17**). Among phenotypes that were not direct measures of depression, the largest genetic correlation effect sizes with MD were with neuroticism (*r_g_* = 0.70, P = 2.02 × 10^-162^) and subjective well-being (*r_g_* = -0.63, P = 1.27 × 10^-26^). Compared to Howard et al (2019), novel findings were restricted to phenotypes with smaller genetic correlations (|*r_g_*| <0.16) likely reflecting the greater power provided by current GWAS summary statistics.

In a phenome wide association study (PheWAS), we first identified associations between MD PGS, health related traits and potentially modifiable environmental factors **(Table S18 & Fig S12)**. We then tested for evidence of possible causal associations between MD and MD-PGC associated traits using bidirectional two-sample Mendelian Randomization (MR) in MR-Base (149 phenotypes; Table S19) and using summary statistics from European-ancestry participants in UK Biobank (256 phenotypes; Table S20), leaving UK Biobank out of the European-ancestry MD GWAS. Lower salt usage, faster walking pace, and higher educational attainment were associated with a reduced liability to MD. Greater body mass index (BMI), bio-impedance trunk adiposity measures, and waist and hip circumference were all associated with an increased risk of MD (absolute effect sizes ranged from 0.031 to 0.109, pFDR ranged from 0.006 to 4.33×10^-15^). A full list of these and other potentially causal associations is shown in **Figure S13** and **Table S19A-C**. MR-Base analyses (**Table S20A- C)**, supported a causal role for BMI and measures of adiposity in conferring liability to MD (absolute effect sizes from 0.021 to 0.089, pFDR from 0.01 to 2.24×10^-5^).

We found evidence of potential causal consequences of MD for a number of health behaviors (e.g., increased alcohol and salt intake, increased TV use, absolute effect sizes ranged from 0.023 to 0.207, pFDR ranged from 0.01 to 8.49×10^-16^) and known disease risk factors (e.g., higher triglycerides, C reactive protein (CRP), and gamma glutamyl transferase levels, lower vitamin D levels, higher diastolic blood pressure and waist circumference, absolute effect sizes ranged from 0.018 to 0.128, pFDR ranged 0.046 to 9.63×10^-22^) **(Figure S13; Table S19D-F)**. Lower gray matter, brain structure volumes, and fluid intelligence score were also implicated as causal consequences of MD (absolute effect sizes from 0.048 to 0.102, pFDR from 0.044 to 3.01×10^-16^, see **Figure S13, Table S20D-F)**.

## Discussion

The current study represents the largest and most-inclusive GWAS of MD to date, providing a yield of 697 independent SNP associations located within 636 independent genetic loci and evidence that neuronal differentiation and receptor clustering are involved in the aetiology of the disorder. 286 high-confidence gene associations were identified (summarized in Table S21) in European ancestries, including *CYP7B1*, a cytochrome P450 enzyme involved in neurosteroid synthesis.

Our results confirm and extend previous findings showing the enrichment of expression signals in excitatory and inhibitory neurons. Importantly, the increased power provided by this genetic analysis provided additional evidence of involvement of amygdala and hippocampal excitatory neurons including granule cells, and medium spiny neurons. The amygdala and hippocampus have been previously implicated from a wide range of human imaging (Whalen et al., 2002; Schmaal et al., 2016) and animal studies of depression (Hall et al., 2001; Jentsch et al., 2002; Warner-Schmidt and Duman, 2006) and medium spiny neurons have also been previously implicated in animal studies of reward and linked to depressive behaviors (Lammel et al., 2014; Soares-Cunha et al., 2020). The enrichment of expression signals in granule cells is of particular interest given the renewal of this cell type throughout adult life in the dentate gyrus (Boldrini et al., 2018), its role in stress resilience (Holland, 2012) and the increased hippocampal granule cell expansion associated with antidepressant treatment (Boldrini et al., 2009). Together, these findings underline the mechanistic insights provided by expansion of GWAS to over half a million depressed individuals.

Lack of ancestral and global diversity remain a significant concern for GWAS, with 86% of studies conducted in participants of European ancestry (Fatumo et al., 2022). Our study included data from 160,611 cases and 1,001,890 controls of non-European diverse ancestries. Unlike most other multi-ancestry GWAS, we used a joint analysis approach and did not exclude individuals with mixed ancestry or ancestry not represented in reference sets. This is becoming ever more important as the number of people with mixed ancestry are increasing in countries such as the USA and the UK (Livingston, 2022). Overall, the additional ancestrally diverse participants helped identify 27 novel genetic associations and enabled for the first time to demonstrate significant genetic risk prediction across diverse ancestry groups.

Using polygenic scores, the proportion of variation in liability to MD explained in European ancestry case control studies also showed a considerable increase from an *R^2^* of 3.2% in our previous analyses, to 5.8% using SbayesR. We also show a significant MD prediction in diverse non-European and admixed ancestries. The SNP-*h^2^* in the current study of 8.4% implies that approximately 69% of the additive genetic variance for MD associated with common SNPs across studies can now be accounted for by polygenic scores. This study provides the first evidence of limited transferability of MD PGS to multiple diverse ancestries, and further emphasizes the importance of conducting future GWAS studies across different global populations, especially in Africa where transferability is poorest. Whilst we did not find evidence for improved prediction based on multi-ancestry rather than European-only PGS, this may be due to the small proportion of participants *within* each individual ancestry group (23% of individuals of Non-European ancestries were divided across 4 major ancestry and admixed groups) relative to the European ancestry group alone.

Causal inference analyses using Mendelian Randomization provide the strongest genetic evidence to date that increased adiposity may increase the risk of MD. We also found that a less favorable profile of many behavioral and biochemical disease risk factors, reduced brain volumes and decreased fluid intelligence may be causal consequences of MD. These findings provide further evidence that behavioral and pharmacological interventions to reduce adiposity may reduce the risk of MD, and that reducing MD risk may have favorable consequences for disease risk, brain health and cognition.

Genome wide association signals for depression also showed enrichment for the targets of antidepressants, suggesting that they may also help to reveal other effective treatment targets and more effective interventions. Pregabalin is an effective treatment for neuropathic pain, focal seizures and generalized anxiety disorder (GAD) (Generoso et al., 2017). Studies have shown that it is effective for the treatment of GAD-related depressive symptoms (Dold et al., 2022), but randomized controlled trial (RCT) evidence supporting its use in MD is weak. While pregabalin is sometimes prescribed to augment other effective treatments in medication-resistant MD (Dold *et al*., 2022), there is now justification to consider its efficacy in MD within the context of an RCT.

Together these findings highlight the value of ancestrally diverse genetic studies to prioritize the study of pathophysiological processes in MD. The clearer enrichment of antidepressant targets provides confidence that genetic association findings will be relevant to the deployment or repurposing of existing treatments. Critically, these findings suggest genetic associations will point to new drug targets and more effective therapies that may reduce the considerable disability caused by depression.

## Supporting information

Full Authorship

## Data Availability

All data produced in the present study are available upon reasonable request to the authors after publication. Data from 23andMe are available upon publication following application to 23andMe.

## Acknowledgements

We would like to thank the participants and investigators from all studies and the research participants and employees of 23andMe for making this meta-analysis possible. Major funding for the PGC is from the US National Institutes of Health (MH124873, MH124871). Statistical analyses were carried out on the NL Genetic Cluster Computer (http://www.geneticcluster.org/) hosted by SURFsara. The iPSYCH team acknowledges funding from the Lundbeck Foundation (grants R102- A9118 and R155-2014-1724), the Stanley Medical Research Institute, the Novo Nordisk Foundation for supporting the Danish National Biobank resource, and the GenomeDK HPC facility. This research has been conducted using the UK Biobank Resource (application 4844) and data from dbGaP (accession phs000021, phs000196, phs000187) and including data from: the Molecular Genetics of Schizophrenia Collaboration (Pablo Gejman, Northwestern University), the NINDS CIDR:NGRC Parkinson’s Disease Study, and the SNP Association Analysis of Melanoma: Case–Control and Outcomes Investigation (supported by FNIH GAIN study, CA093459, CA097007, ES011740, and CA133996). Individual study funding and other acknowledgements are provided in the supplementary study information. This paper represents independent research partly-funded by the NIHR Maudsley Biomedical Research Centre and Maudsley NHS Foundation Trust and King’s College London; the views expressed are those of the authors and not necessarily those of the NIHR or the Department of Health and Social Care. The current work was also supported by the Wellcome Trust (220857/Z/20/Z) and the European Union under the Horizon 2020 research and innovation programme (No 847776 and 948561).

## Code and data availability

Summary statistics are available for download from https://pgc.unc.edu/for-researchers/download-results/ and individual data by application to the Psychiatric Genomics Consortium Data Access Committee https://pgc.unc.edu/for-researchers/data-access-committee/. Available summary statistics including 23andMe data require an approved application to 23andMe here: https://research.23andme.com/dataset-access/. Project code is available from https://github.com/psychiatric-genomics-consortium/mdd-wave3-meta.

