## Supplementary material for "Genome-wide study of half a million individuals with major depression identifies 697 independent associations, infers causal neuronal subtypes and biological targets for novel pharmacotherapies": Full Authorship

Mark J Adams* 1
Fabian Streit* 2, 3, 4
Xiangrui Meng* 5
Swapnil Awasthi* 6
Brett N Adey 7
Karmel W Choi 8, 9
V Kartik Chundru 10, 11
Jonathan RI Coleman 7, 7, 12
Bart Ferwerda 13
Jerome C Foo 2, 14, 15, 16
Zachary F Gerring 17
Olga Giannakopoulou 5
Priya Gupta 18, 19
Alisha S M Hall 2, 20
Arvid Harder 21
David M Howard 7
Christopher Hübel 7, 22, 23
Alex S F Kwong 1, 24
Daniel F Levey 19, 25
Brittany L Mitchell 17
Guiyan Ni 26
Vanessa K Ota 27
Oliver Pain 28
Gita A Pathak 29, 30
Eva C Schulte 31, 32, 33, 34
Xueyi Shen 1
Jackson G Thorp 17
Alicia Walker 26
Shuyang Yao 21
Jian Zeng 26
Johan Zvrskovec 7, 12
Dag Aarsland 35
Ky'Era V Actkins 36
Mazda Adli 6, 37
Esben Agerbo 23, 38, 39
Mareike Aichholzer 40
Allison Aiello 41
Tracy M Air 42
Thomas D Als 39, 43, 44
Evelyn Andersson 45
Till F M Andlauer 46, 47
Volker Arolt 48
Helga Ask 49, 50
Sunita Badola 51
Clive Ballard 52
Karina Banasik 53
Nicholas J Bass 5
Aartjan T F Beekman 54
Sintia Belangero 27
Tim B Bigdeli 55
Elisabeth B Binder 46, 56
Ottar Bjerkeset 57, 58
Gyda Bjornsdottir 59
Julia Boberg 45
Sigrid Børte 60, 61, 62
Emma Bränn 63
Alice Braun 64
Thorsten Brodersen 65
Tanja M Brückl 66
Søren Brunak 53
Mie T Bruun 67
Margit Burmeister 68
Pichit Buspavanich 69, 70
Jonas Bybjerg-Grauholm 71, 72
Enda M Byrne 73
Jianwen Cai 74
Archie Campbell 75, 76
Megan L Campbell 77
Adrian I Campos 78
Enrique Castelao 79
Jorge Cervilla 80, 81
Boris Chaumette 82
Chia-Yen Chen 83
Hsi-Chung Chen 84
Zhengming Chen 85
Sven Cichon 86, 87, 88, 89
Lucía Colodro-Conde 17, 90
Anne Corbett 52
Elizabeth C Corfield 49, 91
Baptiste Couvy-Duchesne 92
Nick Craddock 93
Udo Dannlowski 48
Gail Davies 94
EJC de Geus 95
Ian J Deary 94
Franziska Degenhardt 87, 96
Abbas Dehghan 97, 98
J Raymond DePaulo 99
Michael Deuschle 100
Maria Didriksen 101
Khoa Manh Dinh 102
Nese Direk 103
Srdjan Djurovic 104, 105
Anna R Docherty 106, 107, 108
Katharina Domschke 109
Joseph Dowsett 101
Ole Kristian Drange 57, 110, 111, 112
Erin C Dunn 9, 113
William Eaton 114
Gudmundur Einarsson 59
Thalia C Eley 7
Samar S M Elsheikh 115
Jan Engelmann 116
Michael E Benros 72, 117, 118
Christian Erikstrup 102
Valentina Escott-Price 93
Chiara Fabbri 7, 119
Yu Fang 68
Sarah Finer 120
Josef Frank 2
Robert C Free 121
Linda Gallo 122
He Gao 123
Michael Gill 124
Maria Gilles 100
Fernando S Goes 99
Scott Douglas Gordon 17
Jakob Grove 39, 43, 44, 125
Daniel F Gudbjartsson 59, 126
Blanca Gutierrez 80, 81
Tim Hahn 48
Lynsey S Hall 93
Thomas F Hansen 53, 72, 127
Magnus Haraldsson 128
Catherina A Hartman 129
Alexandra Havdahl 49
Caroline Hayward 130
Stefanie Heilmann-Heimbach 87
Stefan Herms 86, 87
Ian B Hickie 131
Henrik Hjalgrim 132
Jens Hjerling-Leffler 133
Per Hoffmann 86, 87
Georg Homuth 134
Carsten Horn 135
Jouke-Jan Hottenga 95
David M Hougaard 71, 72
Iiris Hovatta 136
Qin Qin Huang 10
Donald Hucks 36
Floris Huider 95
Karen A Hunt 137
Nicholas S Ialongo 114
Marcus Ising 138
Erkki Isometsä 139
Rick Jansen 54
Yunxuan Jiang 140
Ian Jones 93
Lisa A Jones 141
Lina Jonsson 142
Masahiro Kanai 143, 144, 145
Robert Karlsson 21
Siegfried Kasper 146
Kenneth S Kendler 147
Ronald C Kessler 148
Stefan Kloiber 115, 138, 149, 150
James A Knowles 151
Nastassja Koen 77
Julia Kraft 64
Henry R Kranzler 152, 153
Kristi Krebs 154
Theodora Kunovac Kallak 155
Zoltán Kutalik 156, 157, 158
Elisa Lahtela 159
Marilyn Lake 160
Margit Hørup Larsen 101
Eric J Lenze 161
Melissa Lewins 1

Glyn Lewis 5
Liming Li 162, 163
Bochao Danae Lin 164
Kuang Lin 85
Penelope A Lind 17
Yu-Li Liu 165
Donald J MacIntyre 1
Dean F MacKinnon 99
Brion S Maher 114
Wolfgang Maier 166
Victoria S Marshe 115, 167
Gabriela A Martinez-levy 168
Koichi Matsuda 169, 170
Hamdi Mbarek 95
Peter McGuffin 7
Sarah E Medland 17
Susanne Meinert 48, 171
Christina Mikkelsen 101, 172
Susan Mikkelsen 102
Yuri Milaneschi 54
Iona Y Millwood 85
Esther Molina 81, 173
Francis M Mondimore 99
Preben Bo Mortensen 23, 38, 39
Benoit H Mulsant 115, 149
Joonas Naamanka 136
Jake M Najman 174
Matthias Nauck 175, 176
Igor Nenadić 177
Kasper R Nielsen 178
Ilja M Nolte 179
Merete Nordentoft 72, 117, 118
Markus M Nöthen 87
Mette Nyegaard 39, 180, 181, 182
Michael C O'Donovan 93
Asmundur Oddsson 59
Adrielle M Oliveira 183
Catherine M Olsen 184, 185
Hogni Oskarsson 186
Sisse Rye Ostrowski 101, 187
Michael J Owen 93
Richard Packer 188
Teemu Palviainen 159
Pedro M Pan 183
Carlos N Pato 189
Michele T Pato 189
Nancy L Pedersen 21
Ole Birger Pedersen 190
Wouter J Peyrot 54
James B Potash 99
Martin Preisig 79
Michael H Preuss 191, 192
Jorge A Quiroz 193
Miguel E Renteria 194
Charles F Reynolds III 195
John P Rice 161
Saori Sakaue 143, 144, 196
Marcos L Santoro 197
Robert A Schoevers 198, 199
Andrew Schork 39, 200, 201
Thomas G Schulze 2, 32, 99, 202, 203, 204
Tabea S Send 100
Jianxin Shi 205
Engilbert Sigurdsson 128
Kritika Singh 36
Grant C B Sinnamon 206
Lea Sirignano 2
Olav B Smeland 207, 208
Daniel J Smith 209
Tamar Sofer 210
Erik Sørensen 101
Sundararajan Srinivasan 211
Hreinn Stefansson 59
Kari Stefansson 59, 212
Peter Straub 36
Mei-Hsin Su 213
André Tadic 116, 214
Henning Teismann 215
Alexander Teumer 216
Anita Thapar 93, 217
Pippa A Thomson 76
Lise Wegner Thørner 101
Apostolia Topaloudi 218
Shih-Jen Tsai 219
Ioanna Tzoulaki 97, 98, 220
George Uhl 221
André G Uitterlinden 222
Henrik Ullum 101, 223, 224
Daniel Umbricht 225
Robert J Ursano 226
Sandra Van der Auwera 227
Albert M van Hemert 228
Abirami Veluchamy 211
Alexander Viktorin 21
Henry Völzke 216
G Bragi Walters 59
Xiaotong Wang 229
Agaz Wani 230
Myrna M Weissman 231, 232
Jürgen Wellmann 215
David C Whiteman 184
Derek Wildman 230
Gonneke Willemsen 95
Alexander T Williams 188
Bendik S Winsvold 61, 62, 233
Stephanie H Witt 2
Ying Xiong 21
Lea Zillich 2
John-Anker Zwart 60, 61, 62
23andMe Research Team 140
China Kadoorie Biobank Collaborative Group 234
Estonian Biobank Research Team 154
Genes & Health Research Team 235
HUNT All-In Psychiatry 236
The BioBank Japan Project 237
VA Million Veteran Program 238
Ole A Andreassen 207, 208, 239
Bernhard T Baune 240, 241, 242
Klaus Berger 215
Dorret I Boomsma 95, 243
Anders D Børglum 39, 43, 44
Gerome Breen 7, 12
Na Cai 244, 245, 246
Hilary Coon 107
William E Copeland 247
Byron Creese 52
Carlos S Cruz-Fuentes 248
Darina Czamara 66
Lea K Davis 36
Eske M Derks 17
Enrico Domenici 249
Paul Elliott 97, 98, 220, 250
Andreas J Forstner 87, 89, 251
Micha Gawlik 252
Joel Gelernter 19, 29, 253
Hans J Grabe 227
Steven P Hamilton 254
Kristian Hveem 255, 256, 257
Catherine John 188, 258
Jaakko Kaprio 159
Tilo Kircher 177
Marie-Odile Krebs 259
Po-Hsiu Kuo 84, 260
Mikael Landén 21, 142
Kelli Lehto 154
Douglas F Levinson 261
Qingqin S Li 262
Klaus Lieb 116
Ruth J F Loos 191, 263, 264, 265, 266
Yi Lu 21
Susanne Lucae 138
Jurjen J Luykx 164, 267
Hermine HM Maes 213, 268
Patrik K Magnusson 21
Hilary C Martin 10
Nicholas G Martin 17
Andrew McQuillin 5
Christel M Middeldorp 73, 269
Lili Milani 154
Ole Mors 39, 270
Daniel J Müller 115, 149, 150, 271
Bertram Müller-Myhsok 46, 272, 273
Yukinori Okada 143, 274, 275
Albertine J Oldehinkel 129
Sara A Paciga 276
Colin NA Palmer 211
Peristera Paschou 218
Brenda WJH Penninx 54
Roy H Perlis 8, 9, 277
Roseann E Peterson 55

Giorgio Pistis 79
Renato Polimanti 29, 30
David J Porteous 76
Danielle Posthuma 278, 279
Jill A Rabinowitz 189
Ted Reichborn-Kjennerud 49
Andreas Reif 40
Frances Rice 93, 217
Roland Ricken 6
Marcella Rietschel 2
Margarita Rivera 81, 280
Christian Rück 281
Giovanni A Salum 282
Catherine Schaefer 283
Srijan Sen 68, 284
Alessandro Serretti 285
Alkistis Skalkidou 155
Jordan W Smoller 8, 286, 287
Dan J Stein 77
Frederike Stein 288
Murray B Stein 289, 290, 291, 292
Patrick F Sullivan 21
Martin Tesli 293
Thorgeir E Thorgeirsson 59
Henning Tiemeier 294, 295
Nicholas J Timpson 24, 296
Monica Uddin 230
Rudolf Uher 297
David A van Heel 137
Karin JH Verweij 298
Robin G Walters 85
Sylvia Wassertheil-Smoller 299
Jens R Wendland 51
Thomas Werge 72, 200, 223, 300, 301
Aeilko H Zwinderman 13
Karoline Kuchenbaecker* 5, 85
Naomi R Wray* 26, 229, 302
Stephan Ripke* 6, 287
Cathryn M Lewis* 7, 303
Andrew M McIntosh* 1, 304

***** shared first and last authors
V. 2024-04-26 15:00

1, Division of Psychiatry, University of Edinburgh, Edinburgh, UK
2, Department of Genetic Epidemiology in Psychiatry, Central Institute of Mental Health, Medical Faculty Mannheim, Heidelberg University, Mannheim, BW, DE
3, Hector Institute for Artificial Intelligence in Psychiatry, Central Institute of Mental Health, Medical Faculty Mannheim, Heidelberg University, Mannheim, BW, DE
4, Department for Psychiatry and Psychotherapy, Central Institute of Mental Health, Medical Faculty Mannheim, Heidelberg University, Mannheim, BW, DE
5, Division of Psychiatry, University College London, London, UK
6, Department of Psychiatry and Psychotherapy, Charité – Universitätsmedizin Berlin, Berlin, BE, DE
7, Social, Genetic and Developmental Psychiatry Centre, King's College London, London, UK
8, Department of Psychiatry, Massachusetts General Hospital, Boston, MA, US
9, Department of Psychiatry, Harvard Medical School, Boston, MA, US
10, Human Genetics, Wellcome Sanger Institute, Hinxton, UK
11, Department of Clinical and Biomedical Sciences, Faculty of Health and Life Sciences, University of Exeter, Exeter, UK
12, NIHR Maudsley Biomedical Research Centre, King's College London, London, UK
13, Epidemiologie en Data Science (EDS), Amsterdam UMC, location University of Amsterdam, Amsterdam, NL
14, Institute for Psychopharmacology, Central Institute of Mental Health, Medical Faculty Mannheim, Heidelberg University, Mannheim, BW, DE
15, Department of Psychiatry, College of Health Sciences, University of Alberta, Edmonton, AB, CA
16, Neuroscience and Mental Health Institute, University of Alberta, Edmonton, AB, CA
17, Mental Health and Neuroscience, QIMR Berghofer Medical Research Institute, Brisbane, QLD, AU
18, Psychiatry, Yale University, New Haven, CT, US
19, Psychiatry, Veterans Affairs Connecticut Healthcare System, West Haven, CT, US
20, Department of Clinical Medicine, Aarhus University, Aarhus, DK
21, Department of Medical Epidemiology and Biostatistics, Karolinska Institutet, Stockholm, SE
22, Department of Pediatric Neurology, Charité – Universitätsmedizin Berlin, Berlin, BE, DE
23, National Centre for Register-based Research, Aarhus University, Aarhus, DK
24, MRC Integrative Epidemiology Unit, University of Bristol, Bristol, UK
25, Department of Psychiatry, Yale University, New Haven, CT, US
26, Institute for Molecular Bioscience, University of Queensland, Brisbane, QLD, AU
27, Morphology and Genetics, Universidade Federal de Sao Paulo, Sao Paulo, SP, BR
28, Maurice Wohl Clinical Neuroscience Institute, Department of Basic and Clinical Neuroscience, King's College London, London, UK
29, Department of Psychiatry, Yale University School of Medicine, New Haven, CT, US
30, Veterans Affairs Connecticut Healthcare System, West Haven, CT, US
31, Department of Psychiatry and Psychotherapy, University Hospital, LMU Munich, Munich, BY, DE
32, Institute of Psychiatric Phenomics and Genomics, University Hospital, LMU Munich, Munich, BY, DE
33, Department of Psychiatry and Psychotherapy, University Hospital Bonn, Medical Faculty, University of Bonn, Bonn, DE
34, Institute of Human Genetics, University Hospital Bonn, Medical Faculty, University of Bonn, Bonn, DE
35, Old Age Psychiatry, King's College London, London, UK
36, Department of Medicine, Division of Genetic Medicine, Vanderbilt University Medical Center, Nashville, TN, US
37, Department of Psychiatry and Psychotherapy, Fliedner Klinik Berlin, Berlin, BE, DE
38, Centre for Integrated Register-based Research, Aarhus University, Aarhus, DK
39, iPSYCH, The Lundbeck Foundation Initiative for Integrative Psychiatric Research, Aarhus, DK
40, Department of Psychiatry, Psychosomatic Medicine and Psychotherapy, Goethe University Frankfurt - University Hospital, Frankfurt am Main, DE
41, Department of Epidemiology, Columbia University Mailman School of Public Health, New York, NY, US
42, Discipline of Psychiatry, University of Adelaide, Adelaide, SA, AU
43, Department of Biomedicine and Centre for Integrative Sequencing, iSEQ, Aarhus University, Aarhus, DK
44, Center for Genomics and Personalized Medicine, Aarhus University, Aarhus, DK
45, Department of Clinical Neuroscience, Karolinska Institutet, SE
46, Department of Translational Research in Psychiatry, Max Planck Institute of Psychiatry, Munich, BY, DE
47, Department of Neurology, Klinikum rechts der Isar, Technical University of Munich, Munich, BY, DE
48, Institute for Translational Psychiatry, University of Münster, Münster, NRW, DE
49, PsychGen Centre for Genetic Epidemiology and Mental Health, Norwegian Institute of Public Health, Oslo, NO
50, PROMENTA Research Center, Department of Psychology, University of Oslo, Oslo, NO
51, Research and Development, Takeda Pharmaceutical Company Limited, Cambridge, MA, US
52, Faculty of Health and Life Sciences, University of Exeter, Exeter, UK

53, Novo Nordisk Foundation Center for Protein Research, Faculty of Health and Medical Sciences, University of Copenhagen, Copenhagen, DK
54, Department of Psychiatry, Amsterdam Public Health and Amsterdam Neuroscience, Amsterdam UMC, Vrije Universiteit Amsterdam, Amsterdam, NL
55, Department of Psychiatry and Behavioral Sciences, Institute for Genomics in Health, State University of New York Downstate Health Sciences University, Brooklyn, NY, US
56, Department of Psychiatry and Behavioral Sciences, Emory University School of Medicine, Atlanta, GA, US
57, Department of Mental Health, Faculty of Medicine and Health Sciences, Norwegian University of Science and Technology (NTNU), Trondheim, TRD, NO
58, Faculty of Nursing and Health Sciences, NORD University, Levanger, NO
59, deCODE Genetics / Amgen, Reykjavik, IS
60, Institute of Clinical Medicine, Faculty of Medicine, University of Oslo, Oslo, NO
61, K. G. Jebsen Center for Genetic Epidemiology, Department of Public Health and Nursing, Faculty of Medicine and Health Sciences, Norwegian University of Science and Technology (NTNU), Trondheim, TRD, NO
62, Department of Research and Innovation, Division of Clinical Neuroscience, Oslo University Hospital, Oslo, NO
63, Institute of Environmental Medicine, Unit of Integrative Epidemiology, Karolinska Institutet, Stockholm, SE
64, Department of Psychiatry and Psychotherapy, Charité – Universitätsmedizin Berlin, Berlin, DE
65, Department of Clinical Immunology, Roskilde University/Næstved Hospital, Roskilde, DK
66, Department Genes & Environment, Max Planck Institute of Psychiatry, Munich, DE
67, Department of Clinical Immunology, Odense University Hospital, Odense, DK
68, Michigan Neuroscience Institute, University of Michigan, Ann Arbor, MI, US
69, Department of Psychiatry and Psychotherapy, Gender Research in Medicine, Institute of Sexology and Sexual Medicine, Charité – Universitätsmedizin Berlin, Berlin, BE, DE
70, Department of Psychiatry, Psychotherapy and Psychosomatics, Brandenburg Medical School Theodor Fontane, Neuruppin, BB, DE
71, Center for Neonatal Screening, Department for Congenital Disorders, Statens Serum Institut, Copenhagen, DK
72, iPSYCH, The Lundbeck Foundation Initiative for Integrative Psychiatric Research, Copenhagen, DK
73, Child Health Research Centre, University of Queensland, Brisbane, QLD, AU
74, Department of Biostatistics , University of North Carolina at Chapel Hill , Chapel Hill, NC, US
75, Centre for Medical Informatics, Usher Institute, University of Edinburgh, Edinburgh, UK
76, Centre for Genomic & Experimental Medicine, Institute for Genetics and Cancer, University of Edinburgh, Edinburgh, UK
77, SAMRC Unit on Risk & Resilience in Mental Disorders, Department of Psychiatry and Neuroscience Institute, University of Cape Town, Cape Town, SA
78, Statistical genetics, Institute for Molecular Bioscience, The University of Queensland, Brisbane, QLD, AU
79, Department of Psychiatry, Lausanne University Hospital and University of Lausanne, Prilly, VD, CH
80, Department of Psychiatry, Faculty of Medicine and Institute of Neurosciences, Biomedical Research Centre (CIBM), University of Granada, Granada, ES
81, Instituto de Investigación Biosanitaria ibs.GRANADA, Granada, ES
82, Université de Paris Cité, INSERM U1266, Institute of Psychiatry and Neuroscience of Paris, GHU Paris Psychiatry and Neuroscience, Paris, FR
83, Biogen, Cambridge, MA, US
84, Department of Psychiatry, National Taiwan University Hospital, TW
85, Nuffield Department of Population Health, University of Oxford, Oxford, UK
86, Human Genomics Research Group, Department of Biomedicine, University of Basel, Basel, CH
87, Institute of Human Genetics, University of Bonn, School of Medicine & University Hospital Bonn, Bonn, DE
88, Institute of Medical Genetics and Pathology, University Hospital Basel, University of Basel, Basel, CH
89, Institute of Neuroscience and Medicine (INM-1), Research Center Juelich, Juelich, DE
90, School of Psychology, University of Queensland, Brisbane, QLD, AU
91, Nic Waals Institute, Lovisenberg Diakonale Hospital, Oslo, NO
92, Centre for Advanced Imaging, University of Queensland, Saint Lucia, QLD, AU
93, Centre for Neuropsychiatric Genetics and Genomics, Cardiff University, Cardiff, UK
94, The Lothian Birth Cohorts, University of Edinburgh, Edinburgh, UK
95, Department of Biological Psychology & Amsterdam Public Health Research Institute, Vrije Universiteit Amsterdam, Amsterdam, NL
96, Department of Child and Adolescent Psychiatry, Psychosomatics and Psychotherapy, University Hospital Essen, University of Duisburg-Essen, Duisburg, DE
97, MRC Centre for Environment and Health, School of Public Health, Imperial College London, London, UK

98, Imperial College Dementia Research Institute, Imperial College London, London, UK
99, Department of Psychiatry and Behavioral Sciences, Johns Hopkins University School of Medicine, Baltimore, MD, US
100, Department of Psychiatry and Psychotherapy, Research Group Stress Related Disorders, Central Institute of Mental Health, Medical Faculty Mannheim, Heidelberg University, Mannheim, BW, DE
101, Department of Clinical Immunology, Copenhagen University Hospital, Rigshospitalet, Copenhagen, CPH, DK
102, Department of Clinical Immunology, Aarhus University Hospital, Aarhus, DK
103, Department of Psychiatry, Istanbul University, Istanbul, TR
104, Department of Medical Genetics, Oslo University Hospital, Oslo, OSL, NO
105, NORMENT, Department of Clinical Science, University of Bergen, Bergen, NO
106, Virginia Institute for Psychiatric & Behavioral Genetics, Virginia Commonwealth University, Richmond, VA, US
107, Psychiatry Department / Huntsman Mental Health Institute, University of Utah School of Medicine, Salt Lake City, UT, US
108, Center for Genomic Research, University of Utah School of Medicine, Salt Lake City, UT, US
109, Department of Psychiatry and Psychotherapy, Medical Center, University of Freiburg, Faculty of Medicine, University of Freiburg, Freiburg, DE
110, Division of Mental Health Care, St. Olavs Hospital, Trondheim University Hospital, Trondheim, TRD, NO
111, Department of Psychiatry, Sørlandet Hospital, Kristiansand, AG, NO
112, University of Oslo, NORMENT Centre, Institute of Clinical Medicine, Oslo, OSL, NO
113, Center for Genomic Medicine, Massachusetts General Hospital, Boston, MA, US
114, Department of Mental Health, Johns Hopkins, Baltimore, MD, US
115, Centre for Addiction and Mental Health, Toronto, ON, CA
116, Department of Psychiatry and Psychotherapy, University Medical Center of the Johannes Gutenberg University Mainz, Mainz, DE
117, Mental Health Center Copenhagen, Mental Health Services Capital Region of Denmark, Copenhagen, DK
118, Faculty of Health Science, Department of Clinical Medicine, University of Copenhagen, Copenhagen, DK
119, Department of Biomedical and Neuromotor Sciences, University of Bologna, Bologna, IT
120, Wolfson Institute of Population Health, Queen Mary University of London, London, UK
121, School of Computing and Mathematical Sciences, University of Leicester, Leicester, UK
122, Department of Psychology, San Diego San Diego State University, San Diego, CA, US
123, Department of Epidemiology and Biostatistics, Imperial College London, London, UK
124, Discipline of Psychiatry, School of Medicine, Trinity College Dublin, Dublin, IE
125, Bioinformatics Research Centre, Aarhus University, Aarhus, DK
126, School of Engineering, University of Iceland, Reykjavik, IS
127, Danish Headache Centre, Department of Neurology, Rigshospitalet, Glostrup, DK
128, Faculty of Medicine, Department of Psychiatry, University of Iceland, Reykjavik, IS
129, Department of Psychiatry, University of Groningen, University Medical Center Groningen, Groningen, NL
130, MRC Human Genetics Unit, Institute for Genetics and Cancer, University of Edinburgh, Edinburgh, UK
131, Brain and Mind Centre, University of Sydney, Sydney, NSW, AU
132, Department of Epidemiology Research, Statens Serum Institut, Copenhagen, DK
133, Department of Medical Biochemistry and Biophysics, Karolinska Institutet, Stockholm, SE
134, Interfaculty Institute for Genetics and Functional Genomics, Department of Functional Genomics, University Medicine Greifswald, Greifswald, MV, DE
135, Roche Pharmaceutical Research and Early Development, Pharmaceutical Sciences, Roche Innovation Center Basel, F. Hoffmann-La Roche Ltd, Basel, CH
136, SleepWell Research Program and Department of Psychology and Logopedics, University of Helsinki, Helsinki, FI
137, Blizard Institute, Barts and the London School of Medicine and Dentistry, Queen Mary University of London, London, UK
138, Max Planck Institute of Psychiatry, Munich, BY, DE
139, Department of Psychiatry, University of Helsinki, Helsinki, FI
140, 23andMe Research Team, 23andMe, Inc., Sunnyvale, CA, US
141, Department of Psychological Medicine, University of Worcester, Worcester, UK
142, Institution of Neuroscience and Physiology, University of Gothenburg, Gothenburg, SE
143, Department of Statistical Genetics, Osaka University Graduate School of Medicine, Suita, JP
144, Program in Medical and Population Genetics, Broad Institute of Harvard and MIT, Cambridge, MA, US
145, Center for Computational and Integrative Biology, Massachusetts General Hospital, Boston, MA, US
146, Center for Brain Research, Department of Molecular Neuroscience, Medical University of Vienna, Vienna, AT
147, Department of Psychiatry, Virginia Commonwealth University, Richmond, VA, US
148, Health Care Policy, Harvard Medical School, Boston, MA, US

149, Department of Psychiatry, University of Toronto, Toronto, ON, CA
150, Department of Pharmacology & Toxicology, University of Toronto, Toronto, ON, CA
151, Department of Genetics, Rutgers University, Piscataway, NJ, US
152, Department of Psychiatry, Perelman School of Medicine, University of Pennsylvania, Philadelphia, PA, US
153, Mental Illness Research, Education and Clinical Center, Crescenz VA Medical Center, Philadelphia, PA, US
154, Estonian Genome Centre, Institute of Genomics, University of Tartu, Tartu, EE
155, Department of Women's and Children's Health, Uppsala University, Uppsala, SE
156, Department of Epidemiology and Health Systems, Center for Primary Care and Public Health, Lausanne, VD, CH
157, Department of Computational Biology, University of Lausanne, Lausanne, VD, CH
158, Swiss Institute of Bioinformatics, Lausanne, VD, CH
159, Institute for Molecular Medicine Finland - FIMM, University of Helsinki, Helsinki, FI
160, SAMRC Unit on Risk & Resilience in Mental Disorders, Department of Psychiatry and Neuroscience Institute, University of Cape Town, Cape Town, SA
161, Department of Psychiatry, Washington University School of Medicine in St. Louis, St. Louis, MO, US
162, Department of Epidemiology and Biostatistics, School of Public Health, Peking University, Beijing, CN
163, Peking University Center for Public Health and Epidemic Preparedness & Response, Peking University, Beijing, CN
164, Department of Psychiatry and Neuropsychology, School for Mental Health and Neuroscience, Maastricht University Medical Centre, Maastricht, NL
165, Center for Neuropsychiatric Research, National Health Research Institutes, TW
166, Department of Psychiatry and Psychotherapy, University of Bonn, Bonn, DE
167, Center for Translational and Computational Neuroimmunology, Columbia University Medical Center, New York, NY, US
168, Psychiatric Genetics, INPRFM, Mexico City, CDMX, GB
169, Laboratory of Genome Technology, Human Genome Center, Institute of Medical Science, The University of Tokyo, Tokyo, JP
170, Laboratory of Clinical Genome Sequencing, Department of Computational Biology and Medical Sciences, Graduate School of Frontier Sciences, The University of Tokyo, Tokyo, JP
171, Institute for Translational Neuroscience, University of Münster, Münster, NRW, DE
172, Novo Nordisk Foundation Center for Basic Metabolic Research, Faculty of Health and Medical Sciences, University of Copenhagen, Copenhagen, DK
173, Department of Nursing, Faculty of Health Sciences and Institute of Neurosciences, Biomedical Research Centre (CIBM), University of Granada, Granada, ES
174, School of Public Health, University of Queensland, Brisbane, QLD, AU
175, Institute of Clinical Chemistry and Laboratory Medicine, University Medicine Greifswald, Greifswald, MV, DE
176, DZHK (German Centre for Cardiovascular Research), Partner Site Greifswald, Greifswald, MV, DE
177, Department of Psychiatry, University of Marburg, Marburg, DE
178, Department of Clinical Immunology, Aalborg University Hospital, Aalborg, DK
179, Department of Epidemiology, University of Groningen, University Medical Center Groningen, Groningen, NL
180, Centre for Integrative Sequencing, iSEQ, Aarhus University, Aarhus, DK
181, Department of Biomedicine-Human Genetics, Aarhus University, Aarhus, DK
182, Department of Health, Science and Technology, Aalborg University, Aalborg, DK
183, Department of Psychiatry, Universidade Federal de Sao Paulo, Sao Paulo, SP, BR
184, Population Health, QIMR Berghofer Medical Research Institute, Brisbane, QLD, AU
185, The Fraser Institute, Faculty of Medicine, University of Queensland, Brisbane, QLD, AU
186, Humus, Reykjavik, IS
187, Department of Clinical Medicine, University of Copenhagen, Copenhagen, CPH, DK
188, Department of Population Health Sciences, University of Leicester, Leicester, UK
189, Department of Psychiatry, Rutgers University, Piscataway, NJ, US
190, Department of Clinical Immunology, Zealand University Hospital, Køge, DK
191, Charles Bronfman Institute for Personalized Medicine, Icahn School of Medicine at Mount Sinai, New York, NY, US
192, Department of Environmental Medicine and Public Health, Icahn School of Medicine at Mount Sinai, New York, NY, US
193, NMD Pharma, Lexington, MA, US
194, Mental Health and Neuroscience Program, QIMR Berghofer Medical Research Institute, Brisbane, QLD, AU
195, Psychiatry, University of Pittsburgh Medical Centre, Pittsburgh, PA, US
196, Divisions of Genetics and Rheumatology, Department of Medicine, Brigham and Women’s Hospital, Harvard Medical School, Boston, MA, US
197, Department of Biochemistry, Universidade Federal de Sao Paulo, Sao Paulo, SP, BR
198, Department of Psychiatry, University Medical Center Groningen, Groningen, NL

199, Research School of Behavioural and Cognitive Neurosciences (BCN), University of Groningen, Groningen, NL
200, Institute of Biological Psychiatry, Mental Health Center Sct. Hans, Mental Health Services Capital Region of Denmark, Copenhagen, DK
201, Neurogenomics Division, The Translational Genomics Research Institute (TGEN), Phoenix, AZ, US
202, Department of Psychiatry and Psychotherapy, University Medical Center Göttingen, Goettingen, NI, DE
203, Human Genetics Branch, NIMH Division of Intramural Research Programs, Bethesda, MD, US
204, Department of Psychiatry and Behavioral Sciences, SUNY Upstate Medical University, Syracuse, NY, USA
205, Division of Cancer Epidemiology and Genetics, National Cancer Institute, Bethesda, MD, US
206, School of Medicine and Dentistry, James Cook University, Townsville, QLD, AU
207, NORMENT, Institute of Clinical Medicine, University of Oslo, Oslo, OSL, NO
208, Division of Mental Health and Addiction, Oslo University Hospital, Oslo, OSL, NO
209, Center for Clinical Brain Sciences, University of Edinburgh, Edinburgh, UK
210, Beth Israel Deaconess Medica, Harvard Medical School, Boston, MA, US
211, Division of Population Health and Genomics, Ninewells Hospital and School of Medicine, University of Dundee, Dundee, UK
212, Faculty of Medicine, University of Iceland, Reykjavik, IS
213, Virginia Institute for Psychiatric and Behavioral Genetics, Virginia Commonwealth University, Richmond, VA, US
214, Department of Psychiatry, Psychotherapy and Psychosomatics, Dr. Fontheim Mentale Gesundheit, Liebenburg, DE
215, Institute of Epidemiology and Social Medicine, University of Münster, Münster, NRW, DE
216, Institute for Community Medicine, University Medicine Greifswald, Greifswald, MV, DE
217, Wolfson Centre for Young People's Mental Health, Division of Psychological Medicine and Clinical Neurosciences, Cardiff University, Cardiff, UK
218, Department of Biological Sciences, Purdue University, West Lafayette, IN, US
219, Institute of Brain Science & Division of Psychiatry, National Yang-Ming University, TW
220, Imperial College BHF Centre for Research Excellence, Imperial College London, London, UK
221, New Mexico VA Health Care System, Albequerque, NM, US
222, Department of Internal Medicine, Erasmus University Medical Center Rotterdam, Rotterdam, NL
223, Department of Clinical Medicine, University of Copenhagen, Copenhagen, DK
224, Management Section, Statens Serum Institut, Copenhagen, DK
225, Xperimed LLC, Basel, CH
226, Department of Psychiatry, Uniformed Services University of the Health Sciences, Bethesda, MD, US
227, Department of Psychiatry and Psychotherapy, University Medicine Greifswald, Greifswald, MV, DE
228, Department of Psychiatry, Leiden University Medical Center, Leiden, NL
229, Department of Psychiatry, University of Oxford, Oxford, UK
230, Genomics Program, University of South Florida College of Public Health, Tampa, FL, US
231, Division of Epidemiology, New York State Psychiatric Institute, New York, NY, US
232, Department of Psychiatry, Columbia University College of Physicians and Surgeons, New York, NY, US
233, Department of Neurology, Oslo University Hospital, Oslo, NO
234, China Kadoorie Biobank Collaborative Group
235, Genes & Health Research Team
236, HUNT All-In Psychiatry
237, The BioBank Japan Project
238, VA Million Veteran Program
239, KG Jebsen Centre for Neurodevelopmental Research, University of Oslo, Oslo, OSL, NO
240, Department of Psychiatry, University of Münster, Münster, NRW, DE
241, Department of Psychiatry, University of Melbourne, Melbourne, VIC, AU
242, Florey Institute of Neuroscience and Mental Health, University of Melbourne, Melbourne, VIC, AU
243, Department of Complex Trait Genetics, CNCR, Vrije Universiteit Amsterdam, Amsterdam, NL
244, Helmholtz Pioneer Campus, Helmholtz Zentrum München, Neuherberg, DE
245, Computational Health Centre, Helmholtz Zentrum München, Neuherberg, DE
246, School of Medicine, Technical University of Munich, Munich, BY, DE
247, Department of Psychiatry, University of Vermont, Burlington, VT, US
248, Psychiatric Genetics, Instituto Nacional de Psiquiatría Ramón de la Fuente Muñiz (INPRFM), Mexico City, CDMX, MX
249, Department of Cellular, Computational and Integrative Biology, Università degli Studi di Trento, Trento, IT
250, Imperial College Biomedical Research Centre, Imperial College London, London, UK
251, Center for Human Genetics, University of Marburg, Marburg, DE
252, Department of Psychiatry, Psychosomatics and Psychotherapy, Julius-Maximilians-Universität Würzburg, Würzburg, DE

253, Department of Genetics, Department of Neuroscience, Yale University School of Medicine, New Haven, CT, US
254, Psychiatry, Kaiser Permanente Northern California, San Francisco, CA, US
255, K. G. Jebsen Center for Genetic Epidemiology, Department of Public Health and Nursing, Faculty of Medicine and Health Sciences, Norwegian University of Science and Technology (NTNU), Trondheim, NO
256, HUNT Research Center, Department of Public Health and Nursing, Faculty of Medicine and Health Sciences, Norwegian University of Science and Technology (NTNU), Trondheim, NO
257, Department of Research, Innovation and Education, St. Olavs Hospital, Trondheim University Hospital, Trondheim, NO
258, NIHR Leicester Biomedical Research Centre, Glenfield Hospital, Leicester, UK
259, Pathophysiology of Psychiatric Diseases, INSERM, Univ Paris Cité, GHU Paris, Paris, FR
260, Institute of Epidemiology and Preventive Medicine & Department of Public Health, National Taiwan University, TW
261, Department of Psychiatry & Behavioral Sciences, Stanford University, Stanford, CA, US
262, Neuroscience Therapeutic Area, Janssen Research and Development, LLC, Titusville, NJ, US
263, Novo Nordisk Foundation Center for Basic Metabolic Research, Faculty of Health and Medical Science, University of Copenhagen, Copenhagen, DK
264, Mindich Child Health and Development Institute, Icahn School of Medicine at Mount Sinai, New York, NY, US
265, Department of Environmental Medicine and Public Health, Icahn School of Medicine at Mount Sinai, New York, NY, US
266, MRC Metabolic Diseases Unit, University of Cambridge Metabolic Research Laboratories, Wellcome-MRC Institute of Metabolic Science, Addenbrooke's Hospital, Cambridge, UK
267, Second Opinion Outpatient Clinic, GGNet Mental Health, Warnsveld, NL
268, Department of Human and Molecular Genetics, Virginia Commonwealth University, Richmond, VA, US
269, Child and Youth Mental Health Service, Children's Health Queensland Hospital and Health Service, Brisbane, QLD, AU
270, Psychosis Research Unit, Aarhus University Hospital-Psychiatry, Aarhus, DK
271, Department of Psychiatry, Psychosomatics and Psychotherapy, University Hospital of Würzburg, Würzburg, DE
272, Munich Cluster for Systems Neurology (SyNergy), Munich, BY, DE
273, University of Liverpool, Liverpool, UK
274, Department of Genome Informatics, Graduate School of Medicine, The University of Tokyo, Tokyo, JP
275, Laboratory for Systems Genetics, RIKEN Center for Integrative Medical Sciences, Yokohama, JP
276, Human Genetics and Computational Biomedicine, Pfizer Global Research and Development, Groton, CT, US
277, Centre for Quantitative Health, Massachusetts General Hospital, Boston, MA, US
278, Child and Adolescent Psychiatry, Amsterdam UMC, Vrije Universiteit Amsterdam, Amsterdam, NL
279, Complex Trait Genetics, Vrije Universiteit Amsterdam, Amsterdam, NL
280, Department of Biochemistry and Molecular Biology II, Faculty of Pharmacy and Institute of Neurosciences, Biomedical Research Centre (CIBM), University of Granada, Granada, ES
281, Department of Clinical Neuroscience, Karolinska Institutet, Stockholm, SE
282, Psychiatry, Universidade Federal do Rio Grande do Sul, Porto Alegre, BR
283, Division of Research, Kaiser Permanente Northern California, Oakland, CA, US
284, Eisenberg Family Depression Center, University of Michigan, Ann Arbor, MI, US
285, Department of Medicine and Surgery, Kore University of Enna, Enna, IT
286, Psychiatric and Neurodevelopmental Genetics Unit, Massachusetts General Hospital, Boston, MA, US
287, Stanley Center for Psychiatric Research, Broad Institute of MIT and Harvard, Cambridge, MA, US
288, Department of Psychiatry and Psychotherapy, University of Marburg, Marburg, HE, DE
289, Psychiatry and Public Health, University of California, San Diego, La Jolla, CA, US
290, Psychiatry, Veterans Affairs San Diego Healthcare System, San Diego, CA, US
291, Psychiatry, UCSD School of Medicine, La Jolla, CA, US
292, Public Health, UCSD School of Public Health, La Jolla, CA, US
293, Department of Mental Health and Suicide, Norwegian Institute of Public Health, Oslo, NO
294, Child and Adolescent Psychiatry, Erasmus University Medical Center Rotterdam, Rotterdam, NL
295, Social and Behavioral Science, Harvard T.H. Chan School of Public Health, Boston, MA, US
296, Population Health Sciences, Bristol Medical School, University of Bristol, Bristol, UK
297, Psychiatry, Dalhousie University, Halifax, NS, CA
298, Psychiatry, Amsterdam UMC, location University of Amsterdam, Amsterdam, NL
299, Epidemiology and Population Health, Albert Einstein College of Medicine, Bronx, NY, US
300, Institute of Biological Psychiatry, Mental Health Center Sct. Hans, Copenhagen University Hospital, Mental Health Services, Copenhagen, DK
301, GLOBE Institute, Lundbeck Foundation Centre for Geogenetics, University of Copenhagen, Copenhagen, DK
302, Queensland Brain Institute, University of Queensland, Brisbane, QLD, AU

303, Department of Medical & Molecular Genetics, King's College London, London, UK
304, Institute for Genomics and Cancer, University of Edinburgh, Edinburgh, UK
